# LCD Benchmark: Long Clinical Document Benchmark on Mortality Prediction for Language Models

**DOI:** 10.1101/2024.03.26.24304920

**Authors:** WonJin Yoon, Shan Chen, Yanjun Gao, Zhanzhan Zhao, Dmitriy Dligach, Danielle S. Bitterman, Majid Afshar, Timothy Miller

## Abstract

**Objective:** The application of Natural Language Processing (NLP) in the clinical domain is important due to the rich unstructured information in clinical documents, which often remains inaccessible in structured data. When applying NLP methods to a certain domain, the role of benchmark datasets is crucial as benchmark datasets not only guide the selection of best-performing models but also enable the assessment of the reliability of the generated outputs. Despite the recent availability of language models (LMs) capable of longer context, benchmark datasets targeting long clinical document classification tasks are absent.

**Materials and Methods:** To address this issue, we propose LCD benchmark, a benchmark for the task of predicting 30-day out-of-hospital mortality using discharge notes of MIMIC-IV and statewide death data. We evaluated this benchmark dataset using baseline models, from bag-of-words and CNN to instruction-tuned large language models. Additionally, we provide a comprehensive analysis of the model outputs, including manual review and visualization of model weights, to offer insights into their predictive capabilities and limitations.

**Results and Discussion:** Baseline models showed 28.9% for best-performing supervised models and 32.2% for GPT-4 in F1-metrics. Notes in our dataset have a median word count of 1687. Our analysis of the model outputs showed that our dataset is challenging for both models and human experts, but the models can find meaningful signals from the text.

**Conclusion:** We expect our LCD benchmark to be a resource for the development of advanced supervised models, or prompting methods, tailored for clinical text.

The benchmark dataset is available at https://github.com/Machine-Learning-for-Medical-Language/long-clinical-doc

## INTRODUCTION

With the recent emergence of transformer-based Language Models (LMs), research on clinical natural language processing (NLP) has achieved remarkable improvements^1–3^. However, due to the architectural characteristics of transformer models, most available LMs have constraints on the maximum length of the input sequence that a model can process at once, and therefore the majority of available benchmark datasets targets processing of short documents. In the clinical NLP domain, this can be a major technical hurdle for translational applications as the clinical notes can be longer than what most transformer models can process. For example, BERT^4^ and PubMedBERT^5^ models can handle up to 512 tokens at one time, but the discharge summaries in MIMIC-IV have 1,600 words on average, which in token is about six times longer than the 512 token limit.

Recently, LMs capable of longer documents^6,7^ have become available, yet few benchmark datasets to target their ability to process clinical documents are available. These constraints raise the need for long document benchmark datasets to test the ability of developed models and to facilitate the development of models capable of processing longer clinical documents as well.

In this paper, we describe work in developing a benchmark for clinical long document processing models, based on the out-of-hospital mortality prediction task. The source of the dataset is MIMIC-IV v2.2^8^ corpus, specifically discharge notes for patients who were admitted to the ICU and discharged to locations other than hospice facilities. Along with the benchmark dataset, we explore multiple machine learning models for the task, including traditional Support Vector Machine using Bag-of-Words, Convolutional Neural Networks (CNN), a hierarchical transformer encoder^9^, and zero-shot large language models (LLMs) (open-source models and GPT4^6^ via Azure). In the results section, we select three models, the best-performing CNN model, hierarchical transformer, and an open-source instruction-tuned LLM (Mixtral-8×7B-instruct-v0.1^7^) and analyze the outputs. Based on expert physician review, we discovered that the dataset is challenging and at the same time the models can find meaningful signals. We additionally leverage the architecture of the hierarchical transformer model to visualize and quantify the extent to which they jointly consider information from different sections of the discharge summary.

We anticipate that the proposed dataset will serve as a solid foundation for model development and, moreover, as a forum for evaluating LLMs on long clinical document classification tasks^1^. Second, the utilization of predictive models for 30-day mortality at the time of discharge is anticipated to facilitate timely end-of-life discussions with patients and their families. Such conversations are crucial for enhancing the quality of life for patients nearing the end of life, by ensuring that care decisions align with their values and preferences^10–13^.

## MATERIALS AND METHODS

### Medical Information Mart for Intensive Care IV (MIMIC-IV)

The Medical Information Mart for Intensive Care (MIMIC) is a series of publicly available electronic health record (EHR) databases collected from Beth Israel Deaconess Medical Center (BIDMC)^14^. MIMIC databases contain multi-modal data such as text data, structured data (including laboratory data, admission records, and demographic data), and radiograph images for some versions. All the records and text data are de-identified.

MIMIC-IV^8^ is the latest release that encompasses admissions between 2009 and 2019, focused on structured data and text data of ICU patients. We used MIMIC-IV v2.2 data^2^ with discharge summaries and multiple structured records, including out-of-hospital mortality records from Massachusetts State Registry of Vital Records and Statistics^8^.

### Preprocessing

Preprocessing of our benchmark dataset is composed of three steps. First, following the criteria of Harutyunyan et al.^15^, we collected admission records with an ICU stay. In the second step, we merged date of death data using the admission records identifier (*hadm_id*). The third step filtered out records with task-specific restrictions. For our proposed 30-day out-of-hospital mortality prediction dataset, we excluded admissions with in-hospital deaths and admissions where a patient had a discharge disposition of “***hospice***” in structured data because these patients are expected to die shortly after discharge. The training, validation, and testing datasets were partitioned according to patient ID to guarantee that all admissions from the same patient are allocated to the same dataset subset. For the note data, we only utilized discharge notes and not radiology reports. Full details are available in Appendix A, and python implementation of the exact algorithm is available on GitHub repository.

Figure 1 shows the processing flow and Table 1 shows the number of datapoints after processing steps. As shown on the last two rows, our dataset is highly imbalanced; the negative-label notes, which means the patient survived, are about 26 times more abundant than positive-label notes. Of note, the number of admissions in the raw dataset exceeds the number of discharge notes because 99,437 admissions do not have discharge notes.

**Figure 1.**
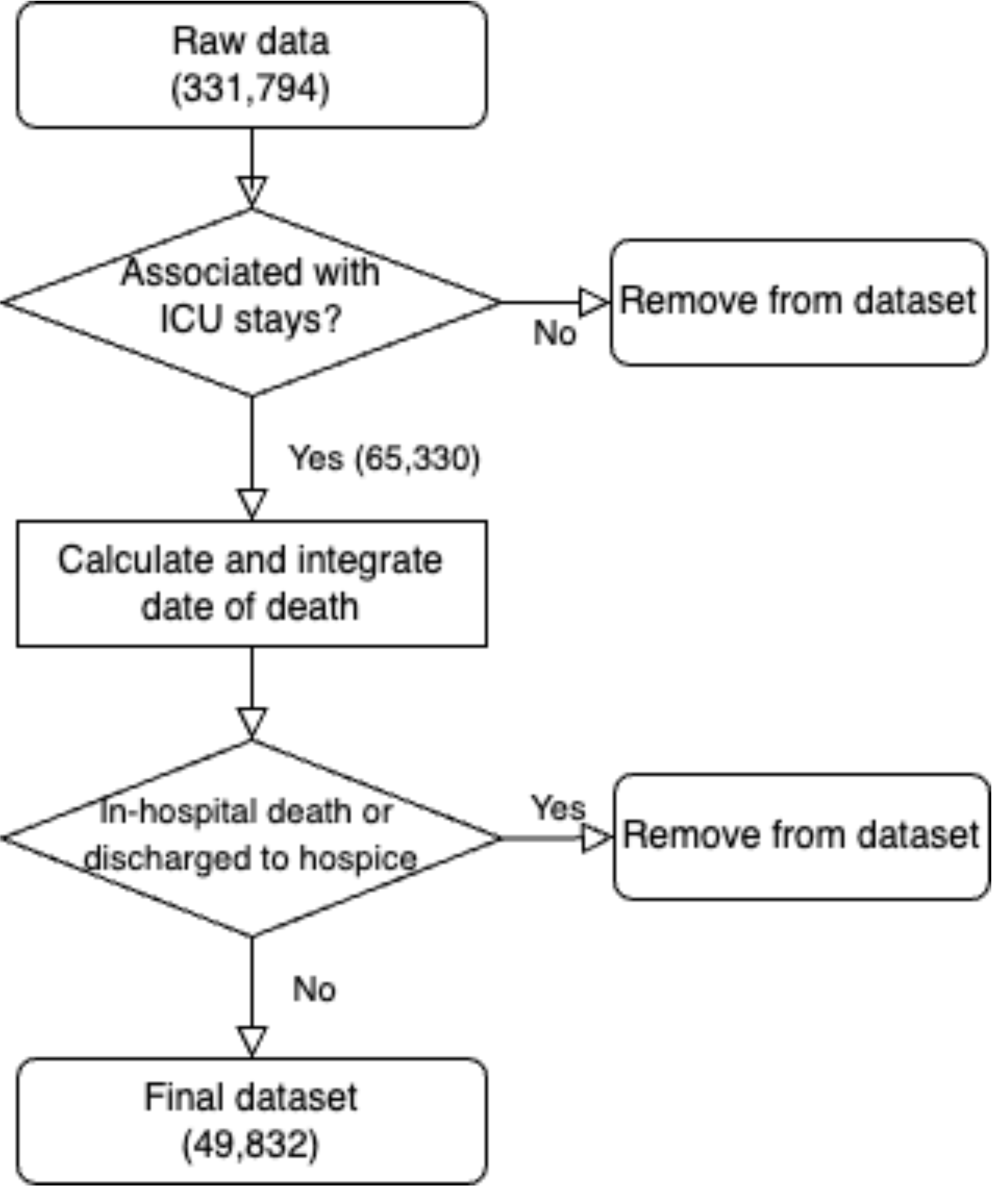
Diagram of the data preprocessing steps. The number of the notes is denoted in the parenthesis.

**Table 1.** Statistics of the datapoints during pre-processing steps. Admission-level data composes a minimum unit of data (often referred as example) and each admission has at most one discharge note. Some patients may have multiple admissions, resulting in several records. These patients might be represented in both the “positive” and “negative” notes categories (asterisked cells).

|  | *# of admissions* | *# of notes* | *# of patients* |
| --- | --- | --- | --- |
| *Raw data* | 431,231 | 331,794 | 180,733 |
| *Data associated with ICU stays* | 65,330 | 65,330 | 50,253 |
| *Final dataset* | 49,832 | 49,832 | 39,705 |
| *-> Positive notes* | 1,830 | 1,830 | 1,772 * |
| *-> Negative notes* | 48,002 | 48,002 | 38,408 * |

Figure 2 displays a histogram of the number of tokens in discharge notes. Each note is tokenized with *microsoft/xtremedistil-l6-h256-uncased* tokenizer and the Hugging Face Transformers library. As this model employs a word-piece tokenizer, a single word can be broken down into subwords and tokenized into multiple tokens depending on the frequency of the word. The median value for the token length were: 3978 (Interquartile range (IQR) 3085 - 5091) for train; 3991 (IQR 3080 - 5103) for development; and 3952 (IQR 3072 - 5072) for test set.

**Figure 2.**
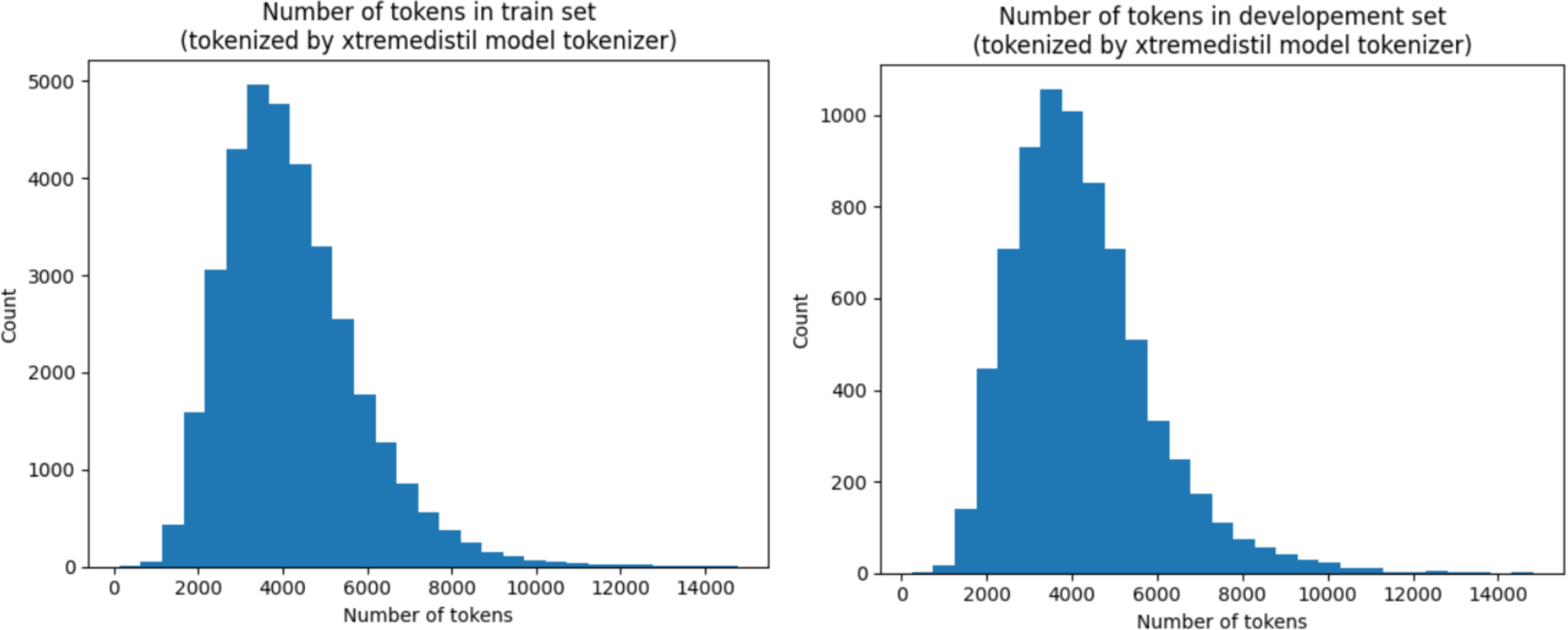
Histogram of the number of tokens in datapoints. Each note in datapoints is tokenized using microsoft/xtremedistil-l6-h256-uncased tokenizer and Huggingface Transformers library. Datapoints in sub-datasets are sorted into 30 bins. Longtail samples that have more than 15,000 tokens are excluded when plotting these graphs.

### Baseline model

#### Bag-of-words (BoW) model

BoW model is a widely used baseline model for NLP where a given text sequence is represented with the frequency of words or word chunks in the sequence. BoW is a strong baseline for document classification tasks with limited training data^16^.

#### Convolutional Neural Networks

Kim et al.^17^ proposed Convolutional Neural Network (CNN) as a feature extractor for the sentence classification task. Our CNN model followed the structure of Kim et al. For complete information about implementation details and hyperparameter settings, please see Appendix B.

### Pretrained transformer models

The transformer is a model architecture that relies on the self-attention mechanism, which is effective at capturing global dependencies within an input sequence. BERT^4^ and GPT^18^ models are some of the early proposed transformer LMs. These models are pre-trained on large-scale corpora and further finetuned to task-specific datasets for supervised learning. Empowered by the pretrained LMs, models tackling clinical NLP tasks have shown remarkable progress.

The self-attention mechanism of early transformer models is implemented by fully connecting each unit of sequence. This requires memory and computational costs that are quadratic with respect to the length of the input sequence, making it a challenge to use transformer models for longer sequences.

#### Longformers

To mitigate this computational limitation for processing long documents, a handful of methods such as blend of local window and global attention approach and sliding window attention ^19–22^ have been proposed.

Longformer^19^ and Clinical-Longformer^20^ are examples of such methods. Clinical-Longformer model was initialized from the pre-trained weights provided by the original authors^3^ and fine-tuned on our dataset.

#### Hierarchical Transformers

Su et al.^9^ introduced a hierarchical transformer, which stack two levels of transformer encoders (Figure 3 - (a)). The hierarchical transformer splits input sequences into smaller chunks and first encodes chunks with a word-level encoder to output chunk representations. The latter part of the structure, chunk-level encoder, works as a feature extractor given the chunk representations of the former part and predicts classes for an input document. Hierarchical transformer models were experimented with two settings, *xtremedistil* model and *PubMedBERT* model as initial weights for the word-level encoder. The chunk-level encoder of the hierarchical transformer model was randomly initialized. Chunk size of hierarchical transformers were tested with two settings, 256 tokens and 512 tokens. In this paper we refer the letter setting as “*Bigchunk*” setting.

**Figure 3.**
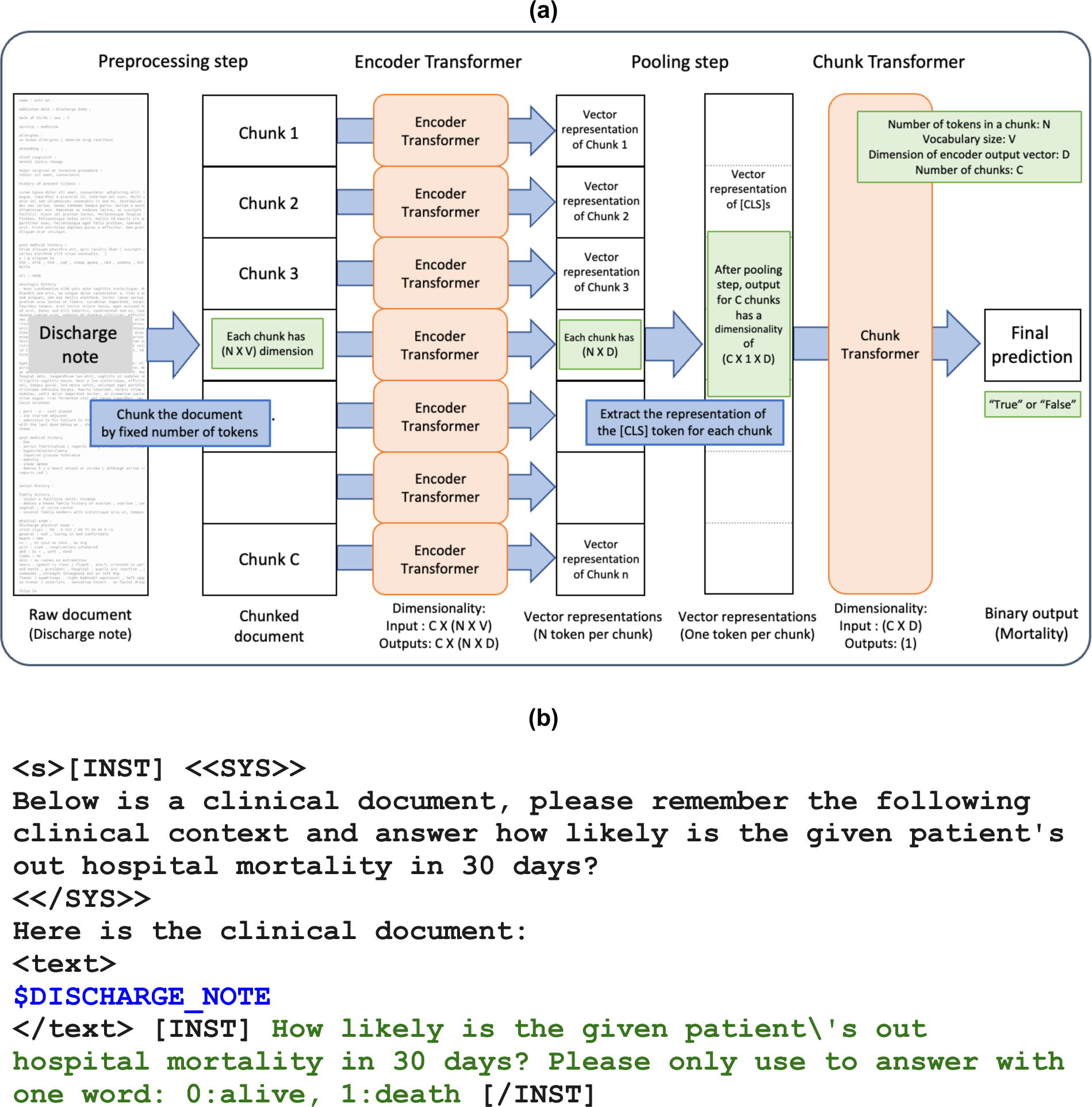
Details about pretrained transformer models, structure and prompts. (a) Structure of hierarchical transformer model. White boxes represent data and orange boxes represent transformer architecture. Green boxes represent dimensionalities of vectors for the step. All Encoder Transformers share weights. The figure shows [CLS] extraction as an example of the “pooling” methods. (b) Our prompt template for LLM experiments. $DISCHARGE_NOTE should be replaced with the actual discharge notes. GPT-4 outputs were generated using this template. Other LLMs utilized the same sentences but with model specific special tokens and templates.

#### LLMs

We explored the ability of zero-shot mortality prediction using LLMs, Mistral (7B-v0.3)^22^, Mixtral (8×7B^4^)^7^, Llama 3 (8B)^23^, Qwen2 (7B, 72B)^24^, Meerkat (7B)^25^, and GPT4-32k^6^ (GPT-4). For the GPT4-32k, we used the HIPAA-compliant version that is provided through Mass General Brigham Azure version 0613. For zero-shot experiments, we used the Hugging Face library to load and inference open-source models and Azure API for the GPT-4 model. These models were selected as they are able to handle context with 32k tokens. Due to hardware limits, a Activation-aware Weight quantized (AWQ)^26^ version was used for the Qwen2 72B model and a maximum token length of 8,192 were applied to all open-sourced models. Figure 3 - (b) shows our *prompting template* for LLMs. The model is asked to choose the answer between *0:alive*, *1:death* and we used a regular expression that looks for the first incidence of “0” or “1” to extract answers from them.

### Experiment details

Our primary metric is F-1 score for the positive labels and we used Receiver Operating Characteristic/Area Under the Curve (ROC AUC) as supplementary metric. Note that we used hinge loss for BoW models, which does not produce probability estimates for the calculation of ROC score. For BoW, CNN and Hierarchical Transformers, we experimented with five or ten runs with identical settings except for the random seeds and averaged the performance to minimize the effect of random initialization of the model.

### Model attention analysis

One of the benefits of the hierarchical transformer model is that it can provide a window into interpretability by highlighting the saliency of each input segment into the model prediction. This becomes possible because the model splits the input into several chunks, each chunk is encoded through an encoder layer, and each encoded chunk representation works as an input unit of the chunk attention layers. By analyzing attention values and the vector norm^27^ of each chunk, we can infer the model’s prioritization of information across various chunks.

Kobayashi et al.^27^ proposed vector norm based analysis, noting that the output vector of each attention layer is a weighted sum of vectors. Following the expression of Kobayashi et al., we denote vector representation of input unit, which is a chunk as we look into chunk-level encoder, at *j*-th position as *x_j_* and attention weight for *j*-th input to *i*-th output unit is denoted as *a_i,j_*. Then, the output vector (*y_i_*) can be expressed as Equation (1) where a function *f(x)* is a simplified notation of value transformation given input unit vector’.

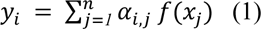

As the equation explains, the output is affected by not only attention weights, *a_ij_* but also transformed input vector, *f(x)*. Norm-based analysis measures the norm of the weighted vector 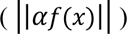 to figure out which input segments are highlighted for a given input sequence. Unlike machine translation tasks where this analysis is first presented, looking into input unit alignment (i.e. finding an input unit that resonates with another word) does not teach us meaningful insights. Rather, we focused on norm of output vector of attention layer, *y_i,_* or 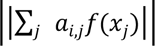 which will directly show the degree of importance of each input unit in the model’s decision.

To investigate the importance of aggregating information across a discharge summary, we use the vector norm method to analyze section importance for this task. We do this by aggregating the two highest vector norm chunks for each instance in the test dataset. Since all inputs have different length, the content in a chunk with a given index can have a different meaning across each sample. Hence, instead of using chunk locations alone, we use section names from chunks for the analysis. The section names were extracted using a rule-based approach.

### Qualitative analysis

For the post-experiment exploratory analysis, we conduct two-step investigations. The first step is dictionary-based detection (i.e. exact match of synonyms list) of mentions about palliative and comfort care measures^5^ and Do Not Resuscitate and Do Not Intubate (DNR/DNI) status. These mentions can be a strong signal for poor prognosis and can be a first filter for data investigation. The second step is to manually review the discharge notes for the left-over samples that do not have such terms. For the manual review, we provide notes, model predictions, true labels, and three questionnaires. Regarding model predictions, predicted binary labels and order of chunk highlights are provided. Labels are set to be hidden by default, and need to click unhide to see the labels. Three questionnaires were “Does this patient label seem valid?”, “Was chunk information useful?”, and “Was this case difficult to predict?”

For comparative analysis, we compare outputs of three models, CNN, Hierarchical Transformer, and Mixtral and manually inspect samples of the benchmark dataset. For this analysis, we focus on open-sourced models for this section as we have more control over the prediction process and the results of these models are more likely to be reproducible.

## RESULTS

Table 2 shows our experimental results for the machine learning models. BoW and CNN models showed strong performance against the fine-tuned transformer models: BoW showed 27.2% F1, CNN showed 28.9% F1. Except for GPT-4, among transformer-based models, hierarchical transformers showed the best performance, which is near the BoW or CNN models. *Bigchunk* model of Hierarchical Transformer models, which refers to chunk size of 512 tokens setting as opposed to normal 256 tokens, showed the best performance of 27.8% F1. Clinical-Longformer showed lower performance when compared with BoW, CNN and hierarchical transformers models regardless of whether the text was truncated from the bottom (right truncated) or top (left truncated) of the document. Mixtral-8×7B-instruct-v0.1 model with zero-shot methods showed performance of 20.5% F1, which is 8% lower than the best performing supervised fine-tuning approach. Our results with GPT-4 showed the best performance of 32.4% F1.

**Table 2.** Performance of models in out-of-hospital mortality prediction task. Our primary metric is the F1 score for positive labels, which are highlighted in bold. Bag-of-words models and LLMs do not generate probability estimates that are necessary for calculating the ROC AUC score. Note that the Max Token column (marked with an asterisk) shows the token settings used in our experiments and does not represent the models’ maximum token settings.

| Method | Model | # Params | Max Tokens * | Negative Label |  |  | Positive Label |  |  | ROC AUC |
| --- | --- | --- | --- | --- | --- | --- | --- | --- | --- | --- |
|  |  |  |  | P | R | F1 | P | R | **F1** |  |
| Sparse vector | Bag-of-Words + SVM |  | - | 0.9720 | 0.9904 | 0.9811 | 0.4431 | 0.2011 | **0.2721** | NA |
| Training from scratch | Convolutional Neural Network | 3 M | 8,192 | 0.9726 | 0.9899 | 0.9812 | 0.4431 | 0.2188 | **0.2899** | 0.8468 |
| Fine-tuning | Hierarchical Transformers (xdistill) | 31 M | 8,192 | 0.9734 | 0.9724 | 0.9729 | 0.2519 | 0.2567 | **0.2526** | 0.8023 |
| Fine-tuning | Hierarchical Transformers (xdistill - bigchunk) | 31 M | 8,192 | 0.9731 | 0.9828 | 0.9779 | 0.3341 | 0.2401 | **0.2788** | 0.8380 |
| Fine-tuning | Hierarchical Transformers (PubMedBERT) | 135 M | 8,192 | 0.9713 | 0.9920 | 0.9816 | 0.4496 | 0.1800 | **0.2566** | 0.8602 |
| Fine-tuning | Hierarchical Transformers (PubMedBERT - bigchunk) | 135 M | 8,192 | 0.9710 | 0.9920 | 0.9814 | 0.4478 | 0.1698 | **0.2418** | 0.8653 |
| Fine-tuning | Clinical Longformer (Right truncate) | 149 M | 4,096 | 0.9694 | 0.9799 | 0.9746 | 0.1923 | 0.1340 | **0.1580** | 0.7362 |
| Fine-tuning | Clinical Longformer (Left truncate) | 149 M | 4,096 | 0.9679 | 0.9767 | 0.9723 | 0.1237 | 0.0919 | **0.1054** | 0.6828 |
| Zero-shot | Qwen2-7B-Instruct | 7B | 8,192 * | 0.9961 | 0.1749 | 0.2976 | 0.0407 | 0.9808 | **0.0782** | NA |
| Zero-shot | Mistral-7B-Instruct-v0.3 | 7B | 8,192 * | 0.9847 | 0.7760 | 0.8680 | 0.0956 | 0.6628 | **0.1671** | NA |
| Zero-shot | Meerkat-7B-v1.0 | 7B | 8,192 * | 0.9741 | 0.937 | 0.9552 | 0.1466 | 0.3027 | **0.1975** | NA |
| Zero-shot | Meta-Llama-3-8B-Instruct | 8B | 8,192 * | 0.9839 | 0.8303 | 0.9006 | 0.1155 | 0.6207 | **0.1948** | NA |
| Zero-shot | Mixtral-8x7B-Instruct-v0.1 | 45B | 8,192 * | 0.9851 | 0.8326 | 0.9025 | 0.1214 | 0.6475 | **0.2045** | NA |
| Zero-shot | Qwen2-72B-Instruct-AWQ | 72B | 8,192 * | 0.9948 | 0.6059 | 0.7531 | 0.0763 | 0.9119 | **0.1409** | NA |
| Zero-shot | GPT-4 (window - 32k) | Unknown | - (32k) | 0.9745 | 0.9839 | 0.9792 | 0.3842 | 0.2797 | **0.3237** | NA |

*BOW = bag-of-words; CNN = convolutional neural network; Params = parameters; P = precision, R = recall, ROC AUC = receiver operating characteristic/area under the curve; SFT = supervised fine-tuned*

### Comparative analysis on model predictions

Figure 4 shows a Venn diagram of the true positive and false positive samples from three models: CNN, Hierarchical Transformer, and Mixtral. The two supervised models have different characteristics when compared to those from zero-shot Mixtral, a LLM. This is unsurprising, as these supervised models are strongly influenced by the dataset models are trained on, whereas the LLMs have presumably never seen the dataset.

**Figure 4.**
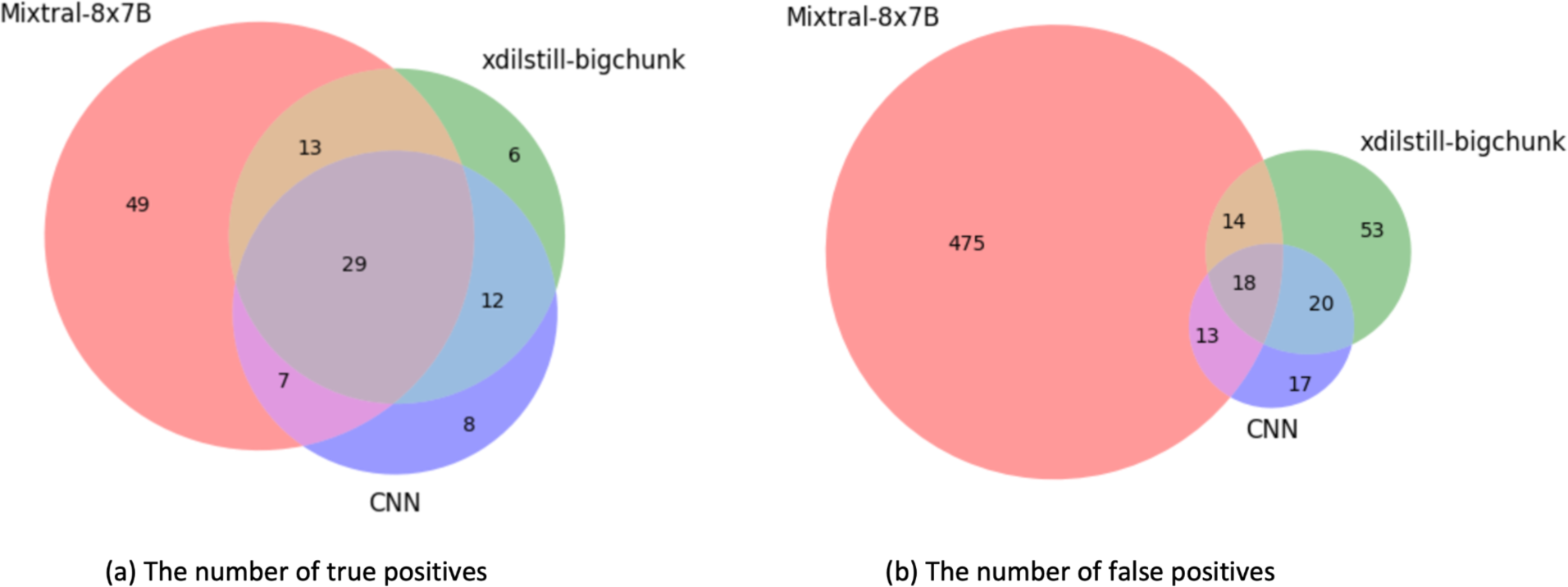
Venn diagram of three model predictions. Numbers in the diagram denote the number of instances in each category. Left diagram (a) shows true positives and the right diagram (b) shows false positive cases.

Sixteen samples were reviewed by a board-certified critical care physician and clinical informatics expert (MA) to understand the face validity of the label and difficulty of the task. The samples were selected across the various categories: three were common true positives, seven were common false positives where three among them were samples without date of death records (For complete information, please see Appendix C). Overall, the physician commented that predicting a specific time window such as 30 or 60 days was difficult. This finding agreed with multiple prior studies showing that prognostication is clinically challenging in patients with serious illness, and even experienced physicians tend to overestimate survival ^28–31^. Incorrect prognostication can hinder end-of-life discussions, lead to more aggressive and potentially over-treatment, and lead to interventions that are not in line with patients’ goals-of-care. In the outpatient oncology setting, machine learning-guided prognostication has been found to improve advanced care planning documentation and serious illness conversations, which could improve end-of-life care. In the inpatient intensive care setting, models such as those developed here could be used to identify patients who may have lower probability of survival to improve end-of-life planning and care.

#### Common predictions

All three models have true positive predictions on 29 instances, which can be considered as easy-to-predict examples. Among 29 instances, 26 are identified as having comfort care mentions in the note and another partition of 26 are identified as having DNR/DNI mentions. All of the 29 instances have at least one of those two keyword sets. Note that patients identifiable as discharged to hospice through structured data were excluded from the dataset during pre-processing steps. Some of the 26 patients had discussion for discharge to *hospice facility* but were not actually discharged to there according to the structured data (cf. they were discharged to home with hospice care or alternative facilities like SKILLED NURSING FACILITY or CHRONIC/LONG TERM ACUTE CARE). The remaining three samples were manually examined. From the structured data, they passed away in 7, 10, and 22 days. Physician analysis was that the labels for these three patient cases had face validity. Based on our analysis, we did not find any anomalies in the labels of all 29 instances.

For false positive predictions, three models have 18 instances in common. Since all machine-learning models predicted these negative instances as positives, instances in this category can be treated as difficult instances. These false positive predictions can be interpreted in multiple ways: the patient’s condition is severely bad but the patient survived, or the prediction is correct but the label is erroneous (please see *Limitation* section for the further discussion). Our dictionary-based detection found comfort care terms from 13 notes and we manually reviewed the rest of five notes where it cannot find the term. Three cases survived less than 1 year and among them, two passed away after 61 and 106 days. One of the other two patients survived about 1 year and 9 months. The last patient did not have a date of death record but our reviewing physician commented that this patient has a high possibility of death in a short period (MIMIC-IV censors death dates at one year after last discharge, so the patient may have survived over one year, or may have been lost to follow-up and died in another state). In summary, some of our data instances raise challenging points to the models, which we believe are important for the discriminative ability of a benchmark dataset. The model predictions were also reasonable and the errors are likely to happen even for a well trained model or domain experts.

#### Distinct predictions

Hierarchical transformer (xdistill-bigchunk) had six distinct true positive predictions that other models failed to predict correctly (Green area in Figure 4 - (a)). These examples can be interpreted as difficult instances as two other models recognized other signals of survival from the text even though they were not correct predictions. This also agrees with the manual analysis, the physician commented that five out of six cases were difficult to predict whether they can survive more than 30 days.

### Attention of Hierarchical Transformer Model

We looked into the vector norm values of the hierarchical transformer to see which chunks, input units of the chunk attention layers, are highlighted during the prediction. Table 3 shows the results of the population-level chunk highlight pairs analysis. The table shows the section combinations and their aggregated frequencies, which shows the summation of section pair weights, normalized by the highest weight. Sections like “Brief Hospital Course” and “Pertinent Results” frequently are in the two most-attended sections.

**Table 3.** Population level section pairs of the most highlighted sections when using hierarchical transformers.

| **Section 1** | **Section 2** | **Adjusted frequency** |
| --- | --- | --- |
| brief hospital course | history of present illness | 1 |
| brief hospital course | admission date : discharge date | 0.882317 |
| brief hospital course | major surgical or invasive procedure | 0.882317 |
| brief hospital course | allergies | 0.882317 |
| brief hospital course | followup instructions | 0.871126 |
| brief hospital course | chief complaint | 0.864856 |
| brief hospital course | name : unit no | 0.819268 |
| history of present illness | pertinent results | 0.74036 |
| brief hospital course | past medical history | 0.714455 |
| history of present illness | followup instructions | 0.711694 |
| major surgical or invasive procedure | pertinent results | 0.691837 |
| allergies | pertinent results | 0.691236 |
| admission date : discharge date | pertinent results | 0.691236 |
| chief complaint | pertinent results | 0.674251 |
| admission date : discharge date | followup instructions | 0.657774 |
| allergies | followup instructions | 0.657774 |
| followup instructions | major surgical or invasive procedure | 0.657774 |
| name : unit no | pertinent results | 0.643258 |
| chief complaint | followup instructions | 0.636643 |
| followup instructions | name : unit no | 0.609727 |

For an in-depth analysis, we looked into prediction of an instance. During prediction of one of the notes without comfort care mentions, the model had highlights on the 5th chunk that has *# icu course* part of *brief hospital course:* and the last chunk, which has discharge information where a part of *discharge medications:, discharge disposition:, discharge diagnosis:, discharge condition:, and discharge instructions:,* and *discharge instructions:* sections are written (Figure 5). The brief hospital course provides informative background about the clinical findings pertaining to a patient’s brain injury, while the discharge information provides complementary, non-overlapping information indicating the level of severity of the injury and mental status at the time of discharge.

**Figure 5.**
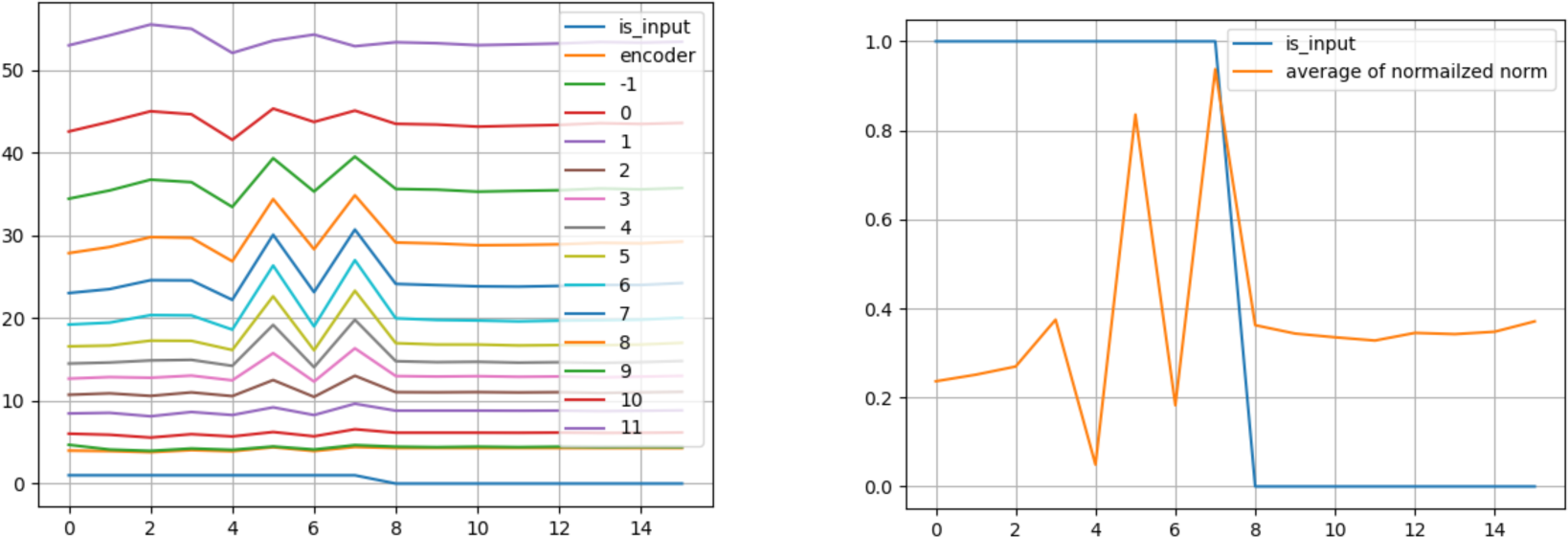
Model highlights during the prediction of an example instance. The position of the chunk is represented on the X-axis, and the vector norm of the layers is shown on the Y-axis. The left graph shows all layers and the right graph shows the average of all layers. Value of “is_input” denotes whether the chunk is composed of actual inputs versus padding tokens. In this graph, chunks in the first to 7th position have real input values but from the 8th chunk, chunks are filled with padding tokens.

a. of Table 4 is a part of the 5th chunk. In this chunk, we note the patient has evidence of hypoxic brain injury and remained in a non-cognitive state that required dependence on breathing and feeding life support.
b. of Table 4 is a part of the last chunk. In this chunk, we could again confirm that the patient had hypoxic ischemic brain injury and low blood sugars, while gaining new information about her mental status and clinical condition at the time she was discharged. While it is reasonably clear from the latter section that the patient’s condition has a poor prognosis, the earlier section contains detailed information of their problems that could give the model more fine-grained information that could modulate the model’s estimation of their condition’s severity and neurologic function.

**Table 4.**
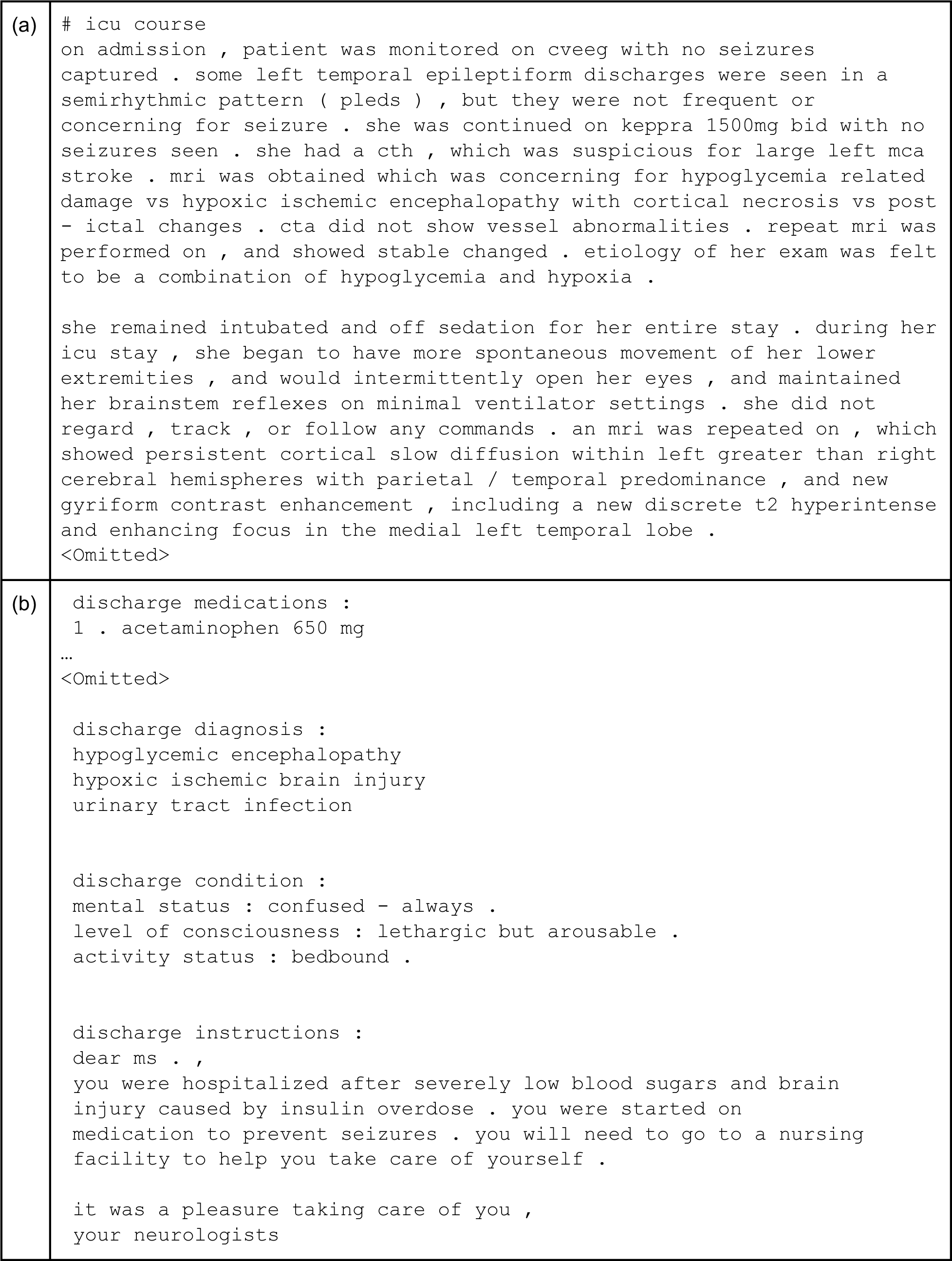
The 5th chunk (a) and the last chunk (b) of the example analyzed in the main text.

## Discussion

During our experiment, significant discrepancies between precision and recall scores were observed for open-sourced large language models (LLMs), suggesting that the label distribution of the predictions does not align well with that of the benchmark dataset. We examined the predictions of open-source LLMs and found that the proportion of positive labels severely differed from the true labels. In other words, only 3.45% of the notes in the test dataset were positively labeled. However, predictions by open-source LLMs varied widely, ranging from 7.12% by Meerkat to 83% by Qwen2 (More details available in Appendix D). The performance and the difference in ratios exhibited a strong negative correlation. We interpret these observations as reasonable, given the inherent difficulties of a zero-shot setting where, unlike in supervised approaches, the model cannot learn the distribution from the original dataset.

MIMIC-IV v2.2 dataset utilized the Massachusetts Registry of Vital Records (cf. Death Certificate is public record in the state of Massachusetts) to enrich the date of death record. According to the MIMIC-IV paper, the state registry was selected instead of the Social Security Death Master File due to data quality concerns^32^. However using the state registry cannot fully resolve the data concerns as patients who moved out of the state cannot be traced with this method. For example, among 18 instances of common false positive cases (i.e. union of three models used in the Comparative analysis section), three patients do not have date of death (DoD) records. We requested the physician expert to review these instances and found out that all of these patients are severely ill and less likely to survive long enough after discharge, meaning that these three labels may be erroneous. Despite this intrinsic limitation, we believe the state registry is still one of the most viable options when creating a database.

### Clinical Impacts and Ethical Considerations

Our study proposes a benchmark dataset that can facilitate development of Language Models to predict 30-day mortality risks using discharge notes. Developing accurate AI models for prognostic prediction offers several potential benefits such as patients’ emotional well-being and proper care planning. For example, for low-risk patients, it could reduce unnecessary care, while providing doctors with valuable references to improve health outcomes for high-risk patients. Additionally, it aids in preparing extremely high-risk patients and their families for end-of-life care and assists hospitals and policymakers in prioritizing resources during health crises.

Despite the importance of having accurate estimates of patient outcomes, studies have shown that such predictions are difficult for both clinicians^33^ and patients^34^, which can lead to disparity between end-of-life (EOL) preferences and actual EOL treatment^35^. A study on AI-based prediction predictions^36^ shows that both patients and physicians answered positively in the interview about an option of AI prognosis model provided. According to the study, both parties supported use of AI models in clinics.

However, no predictive model of mortality will ever be perfect, and classifier errors have the potential to cause significant ethical concerns^36^. Misuse without a thorough understanding of its limitations could lead to, for example, undertreatment by physicians, or negative impact to patient decision-making. While mitigating these concerns is largely a social effort, our future work will also investigate technical approaches, including developing agent-based models that simulate decision-making. This model would simulate interactions between patients, doctors, and policymakers to study how their understanding and interpretation of the mortality prediction model affects health outcomes. Analyzing these dynamics can contribute empirical results to the societal discussion about predictive models, and ensure the responsible incorporation of predictive models into healthcare, balancing innovation with ethical considerations and patient safety.

## CONCLUSION

In this paper, we present a benchmark for evaluating long clinical document processing, entitled LCD benchmark. We tested our benchmark dataset using baseline methods, ranging from Bag-of-words to zero-shot prediction with LLMs. As a result of these methods along with further analysis, we showed that the LCD benchmark presents challenges and the potential for improvement in current neural network-based approaches. During our experiments with LLMs, we further explored the importance of their capability to process longer sequences. Our benchmark dataset is publicly available for the researchers who gained access to the MIMIC-IV datasets and the results can be shared with the CodaBench platform^37^.

## DATA AVAILABILITY STATEMENT

The data underlying this article are available in a github repository, at https://github.com/Machine-Learning-for-Medical-Language/long-clinical-doc

The datasets were derived from sources: https://physionet.org/content/mimiciv/2.2/ and https://physionet.org/content/mimic-iv-note/2.2/

## AUTHOR NOTE

For the large language models we used, we do not have control of their training materials. Our experiments, including those with LLMs, were conducted in HIPAA Protected environments, which blocks third parties from using our data for reviewing or training purposes.

## Author contributions

WY, SC, YG, DD, DB, MA, and TM conceptualized the research, developed methodology. WY conducted data curation, formal analysis and visualization. WY and SC did investigation and provided software for the experiment. MA conducted manual analysis of the samples. TM acquired the funding, provided supervision, and administered the project. WY, SC, ZZ, DB, MA and TM drafted the original manuscript. WY and TM verified the data. All authors were involved in writing - review and editing and approved the manuscript.

## Acknowledgements

We would like to thank Hyunjae Kim of Korea University for providing feedback on the experimental code.

## APPENDIX for LCD Benchmark: Long Clinical Document Benchmark on Mortality Prediction for Language Models

A. Preprocessing details
B. Data selection

In our dataset, each admission record, uniquely identified by the key “hadm_id”, serves as a datapoint. Although MIMIC-IV includes both discharge notes and radiology reports, our study only focuses on discharge notes. Hence, any reference to “notes” throughout this paper denotes discharge notes.

Following the criteria of Harutyunyan et al.^1^, we collected admission records with an ICU stay. Original MIMIV-IV dataset v2.2 has 431,231 admission records and 331,794 discharge notes from 180,733 patients. Among the admissions, only 65,330 have ICU stay records. Some admissions may not have associated discharge notes (Table 1). However, when an admission does have a note, it is always just one discharge note per admission. For admissions that include an ICU stay, each one has an associated discharge note. This means that for the final benchmark dataset, a unit of datapoint is about an admission record with a discharge note and a label.

After the initial data selection, we applied additional task-specific restrictions, resulting in 49,832 notes forming the final dataset. During this phase, we excluded admissions that ended in in-hospital deaths and those with a discharge disposition of “hospice” noted in the structured data. The rationale for these exclusions is that these patients are expected to die shortly after discharge.

a. Label creation

Our label for the out-of-hospital mortality task is calculated based on the *dischtime* record in ***admissions.csv*** and *dod* (abbreviation for date of death) in ***patients.csv*** of MIMIC-IV dataset (note that we used *dod* instead of *deathtime* in ***admissions.csv*** as *deathtime* only includes in-hospital-death). Our threshold for the label is 30 days (inclusive) and the positive label means the patient died within 30 days from the discharge date. We do not count the specific time of day in this calculation. For example, if a patient passes away the next day, we count the time delta as *one full day* regardless of the time of day. This is due to the nature of *dod* records, which records date of death only.

a. Text cleaning

For the note data, we only utilized discharge notes and not radiology reports. New line characters and horizontal tabulation (\t) in the note were replaced with and a space, respectively.

**Table A1.**
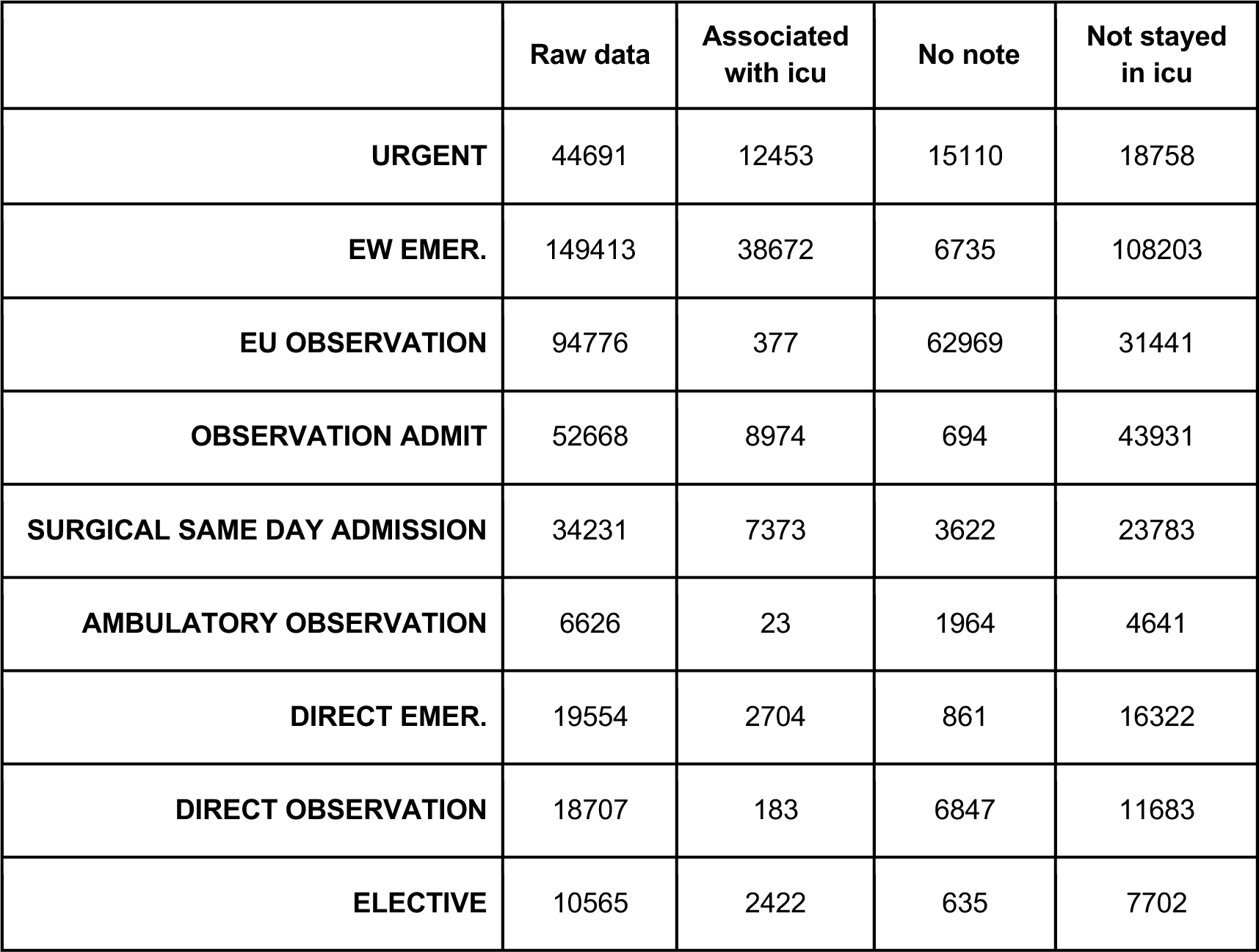
Statistics by MIMIC-IV admission types. Numbers represent the admissions for each types.

A. Implementation details and Hyperparameter settings
B. Bag-of-words models were implemented using scikit-learn^1^. CNN, Hierarchical transformers, and Clinical-Longformer models were trained and tested on the CNLPT library ^2^ (available on GitHub: https://github.com/Machine-Learning-for-Medical-Language/cnlp_transformers). The models were evaluated against the dev set during the training time, and the best performing checkpoints were selected based on the average of Accuracy and the F-1 score.

CNN model and hierarchical transformer models have flexibility in selecting the maximum sequence length (max_seq_length), as unlike most language models, these models can expand the window without pre-training again from scratch.

We selected max_seq_length to be 8192 tokens, which can cover 97% of the notes in the train and development set without truncation (based on xtremedistil model tokenizer).

Since the open-sourced Clinical-Longformer only supports maximum sequence length of 4096, we tested both right-truncation and left-truncation settings, i.e. truncating the ending part and the beginning part of the input sequence respectively.

a. Bag-of-Words (BoW):
b. Backgrounds: BoW models learn vocabulary occurrence information but do not utilize the information of the order of word chunks in an input. Hence, they have a very limited ability to use syntactic information. The size of the word chunk, which could be one or a few words depending on the window size, can add the ability to represent local syntactic information, but it can also make vocabularies sparse and very large. Despite these limitations, BoW is a strong baseline for document classification tasks with limited training dataset.
c. Hyperparameter search for the BoW model was only performed on the n-gram window of the vectorizer, and we selected best performing settings based on experiments on the development dataset, which was using unigram and bigram.

CountVectorizer module with monogram and bigram and SGDClassifier with default settings were used (hinge loss, max_iter=1000, tol=1e-3).

a. CNN:
b. Our CNN model implementation followed the structure of Kim et al.^3^ with minor differences on embedding layer and hyperparameter settings: Kim et al. used word vectors pre-trained with continuous bag-of-words architecture namely word2vec (Mikolov et al.^4^), whereas our model used a randomly initialized embedding layer of 100 dimensions.
c. Learning rate: 2e-6

Batch_size: 4

CNN_num_filters: 500

Warmup steps:5000

Max epochs: 100

Max_seq_len: 8192

a. Hierarchical transformers:
b. Xdistill

Chunk_len: 256

Number of chunks: 32

Learning rate: 2e-6

Batch_size: 4

Layer (Chunk encoder): 12

Warmup steps:5000

Max epochs: 100

Max_seq_len: 8192

i. xdistill-bigchunk

Chunk_len: 512

Number of chunks: 16

Learning rate: 2e-6

Batch_size: 4

Layer (Chunk encoder): 12

Warmup steps:5000

Max epochs: 100

Max_seq_len: 8192

a. Longformer: Learning rate: 2e-5

Batch_size: 4 (1*4 Gradient accumulation)

Warmup steps:5000

Max epochs: 100

Max_seq_len: 4096

A. Error analysis selection
B. Following are the characteristics of the 16 samples we reviewed.

TP refers to True Positive; FP refers to False Positive; ComfortPos means comfort care mentions are found; ComfortNeg means comfort care mentions are not found; DOD_Nan means do not have ‘dod’ records; Unique-hier means that among three models, only hierarchical transformer predicted correctly

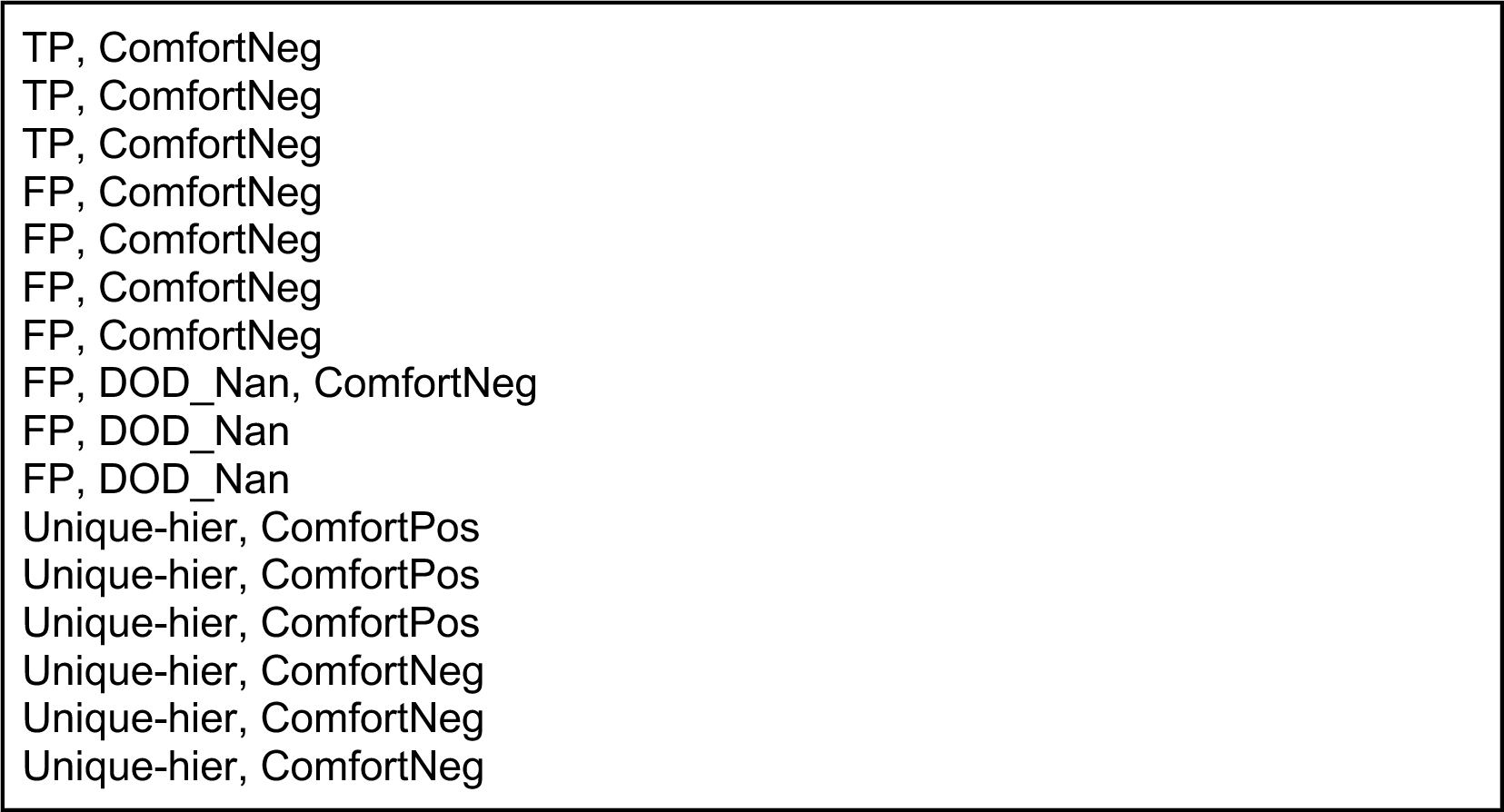

A. Label distribution of the LLM predictions
B. Table 2 illustrates the distribution of positive and negative labels in the predictions made by the Large Language Model (LLM). We visualized the “Positive F1” column from the table (y-axis) and the difference between the actual distribution of “Pos/Total” in true labels (3.45%) and model predictions (x-axis) in Figure 1. From the figure, we can observe that these two variables have a negative correlation.
C. Limitation: Post-hoc experiments - different settings in baseline models
D. Models inherently have different settings due to the nature of their architectures. One of the notable setting differences is the variance in maximum token length for input instance across the models.

**Figure A1.**
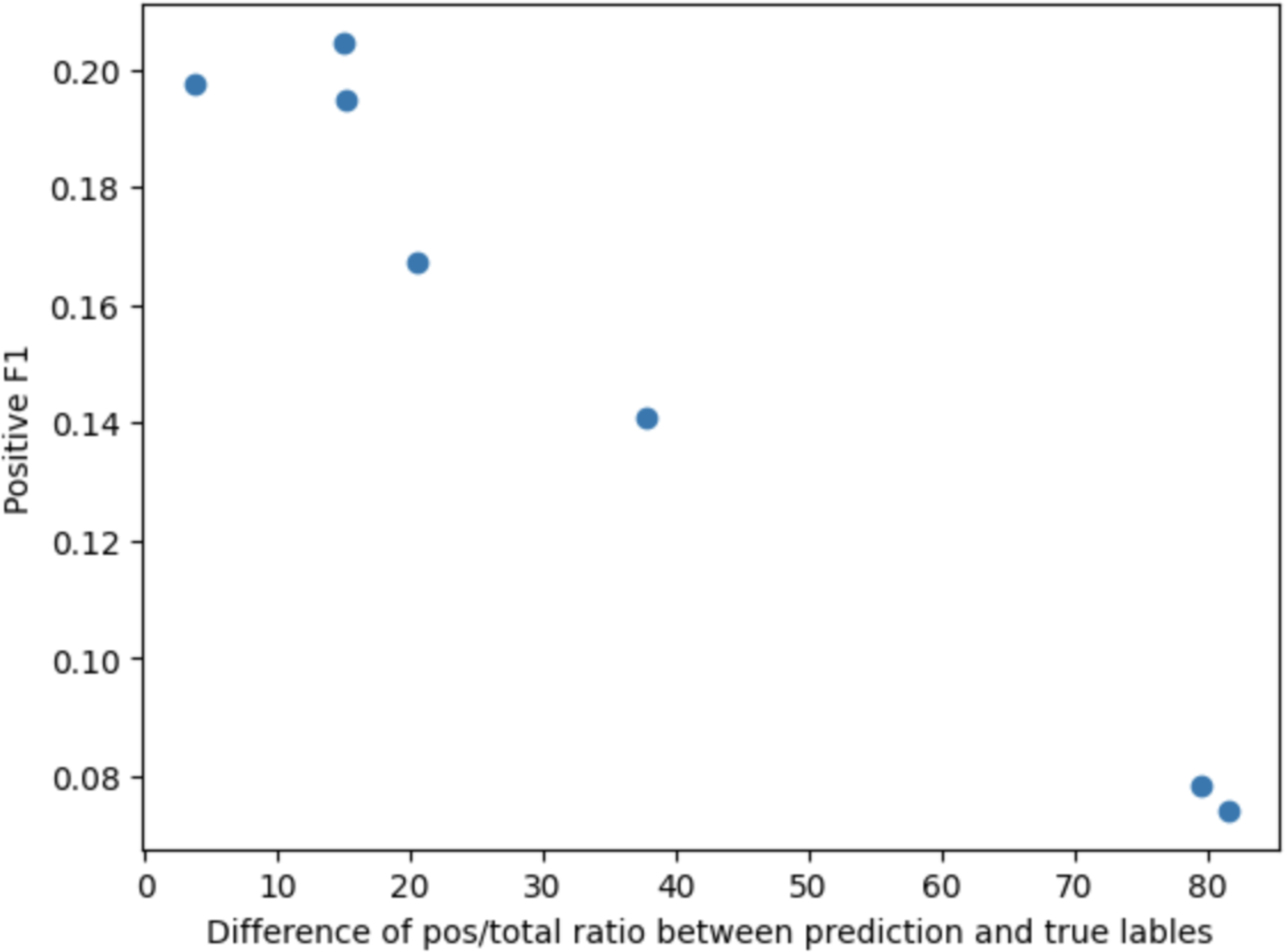
Scatter plot of Positive label F1 score (y-axis) and the difference between the actual distribution of “Pos/Total” in true labels (3.45%) and model predictions (x-axis).

**Table A2.**
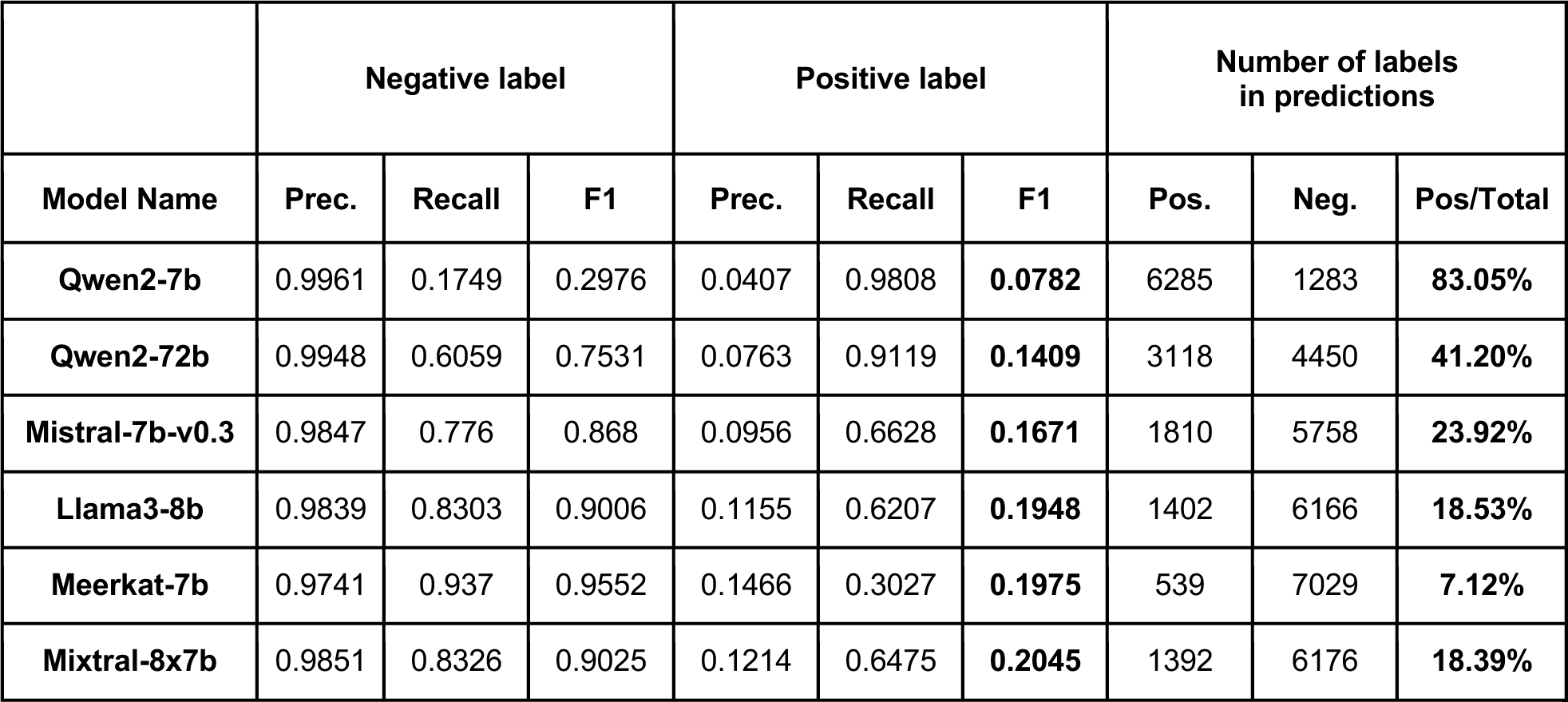
The distribution of positive and negative labels in the predictions made by the Large Language Model. Performances of the zero-shot predictions are given as a reference.

Max token length can be more impactful for large LMs. Prompts for large LMs include system prompts, questions, and the input sequences.

Post-hoc experiments on Mixtral showed that when the maximum token length is limited to 2048, the performance dropped by 11 percent in absolute difference, which is about half of the performance of the full-length model (Table 3).

a. For the zero-shot setting with large LMs, the performance of the models relies on how the prompt is formulated. Sometimes the model cannot produce answers that comply with the suggested answer format. For example, our prompt requires the model to answer only between 0:alive or 1:death but sometimes answers were started with “Based on the information provided,” not matching the requested format. 165 predictions from Mixtral 8*7B included the above mentioned phrase. To alleviate this problem, Gao et al.^5^ proposed an alternative prompt method for zero-shot evaluation named harness. However, this approach can only be applied to models that support output of probability, meaning that most cloud-based models like GPT-4 cannot be evaluated using this method.
b. Note: The Mixtral outputs used in analysis in Section C and E were generated using the exact prompt provided in the main manuscript. Unlike the results presented in the performance table in the main article, these outputs were not produced using the apply_chat_template function.

**Table A3.** Performance of Mixtral* model by the length of input text.

|  | Performance |  |  | Predictions |  |  |
| --- | --- | --- | --- | --- | --- | --- |
| Length | Prec | Rec | F1 | Positive | Negative | Pos/Neg |
| 2048 | 0.08 | 0.17 | 0.11 | 560 | 7008 | 7.40% |
| 8196 | 0.16 | 0.38 | 0.22 | 618 | 6950 | 8.17% |

## Footnotes

1 The benchmark dataset and leaderboard are available at https://github.com/Machine-Learning-for-Medical-Language/long-clinical-doc and https://www.codabench.org/competitions/2064/

2 Published: Jan. 6, 2023. https://physionet.org/content/mimiciv/2.2/

3 https://huggingface.co/yikuan8/Clinical-Longformer

4 Mixtral-8×7B-Instruct-v0.1

5 Comfort care term list: “hospice”, “comfort measures”, “comfort care”, “palliative care”

